# Improving Tuberculosis Detection in Chest X-ray Images through Transfer Learning and Deep Learning: A Comparative Study of CNN Architectures

**DOI:** 10.1101/2024.08.02.24311396

**Authors:** Alex Mirugwe, Lillian Tamale, Juwa Nyirenda

**Affiliations:** School of Public Health, Makerere University, Kampala, Uganda; Department of Computing and Information Science, Victoria University, Kampala, Uganda; Department of Statistical Science, University of Cape Town, Cape Town, South Africa

**Keywords:** Tuberculosis Detection, Chest X-ray Classification, Convolutional Neural Networks, Data Augmentation, Medical Image Analysis

## Abstract

**Introduction:** Tuberculosis remains a significant global health challenge, necessitating more efficient and accurate diagnostic methods.

**Methods:** This study evaluates the performance of various convolutional neural network (CNN) architectures— VGG16, VGG19, ResNet50, ResNet101, ResNet152, and Inception-ResNet-V2—in classifying chest X-ray (CXR) images as either normal or TB-positive. The dataset comprised 4,200 CXR images, with 700 labeled as TB-positive and 3,500 as normal. We also examined the impact of data augmentation on model performance and analyzed the training times and the number of parameters for each architecture.

**Results:** Our results showed that VGG16 outperformed the other models across all evaluation metrics, achieving an accuracy of 99.4%, precision of 97.9%, recall of 98.6%, F1-score of 98.3%, and AUC-ROC of 98.25%. Surprisingly, data augmentation did not improve performance, suggesting that the original dataset’s diversity was sufficient. Furthermore, models with large numbers of parameters, such as ResNet152 and Inception-ResNet-V2, required longer training times without yielding proportionally better performance.

**Discussion:** These findings highlight the importance of selecting the appropriate model architecture based on task-specific requirements. While more complex models with larger parameter counts may seem advantageous, they do not necessarily offer superior performance and often come with increased computational costs.

**Conclusion:** The study demonstrates the potential of simpler models such as VGG16 to effectively diagnose TB from CXR images, providing a balance between performance and computational efficiency. This insight can guide future research and practical implementations in medical image classification.

## INTRODUCTION

Tuberculosis (TB) remains one of the leading infectious diseases worldwide, affecting an estimated one-third to one-fourth of the global population with the *bacillus Mycobacterium tuberculosis*, the causative agent of TB [1]. In 2019, it was estimated that over 10 million individuals globally contracted TB, yet only 71% were detected, diagnosed, and reported through various countries’ national TB programs, leaving approximately 29% of cases unreported [2]. According to the World Health Organization’s (WHO) 2023 TB report, TB was identified as the second most common cause of death among infectious diseases [3]. Furthermore, the global incidence rate of TB remains alarmingly high at approximately 133 new cases per 100,000 people annually [3]. This situation underscores the need for prompt, effective, and affordable screening and treatment strategies to meet the WHO’s ambitious goals of reducing TB incidence by 80%, decreasing TB mortality by 90%, and eliminating catastrophic financial burdens on families affected by TB by 2030 [4].

The World Health Organization (WHO) advised member countries to proactively conduct TB screenings and detection, especially within the high-risk groups, taking into account their unique epidemic scenarios and financial levels [5]. While bacteriological tests, including sputum cultures, sputum smears, and molecular diagnostics, are considered the gold standard for identifying active TB cases, their applicability on a large scale, particularly among high-risk populations, is not feasible [6]. This limitation is due to the methods being resource-intensive, logistically challenging, and associated with prolonged turnaround times [7]. As a result, chest radiography has become the most prevalent method for early TB detection [8]. However, in countries with limited resources, which also bear the highest TB burden, the availability of chest radiography screenings remains inadequate, primarily due to a shortage of radiologists [6].

In recent years, significant advancements have been made in leveraging artificial intelligence, particularly through machine learning and deep learning techniques, for analyzing chest X-ray (CXR) images to differentiate between TB-positive and TB-negative images [9-15]. This innovation has enabled individuals without radiology expertise to conduct TB screening tests, presenting a significant shift in diagnostic approaches. These technologies have shown promising results, to the extent of outperforming radiologists in the interpretation of CXR images [14, 15]. In this research, we investigate the effectiveness of different convolutional neural network (CNN) architectures in classifying tuberculosis in CXR images. We compare and evaluate the performance of popular CNN models including ResNet, Inception, and VGG, and examine the impact of different hyperparameters on classification accuracy. To the best of our knowledge, this study is the first to utilize a larger and more diverse dataset and conduct a comprehensive comparison of the latest CNN architectures, including ResNet101, ResNet152, and Inception-V2, assessed across different parameters. The research aims to address the following questions

1. How does the choice of CNN architecture affect the classification performance?
2. What is the optimal hyperparameter configuration for each CNN architecture?
3. Can transfer learning be leveraged to improve classification accuracy?
4. How does incorporating data augmentation techniques impact the model’s performance compared to training solely on real images?

The rest of the paper is organised as follows. In the next section, we present the literature review, which provides an overview of the current state of research in the field. This is followed by the methodology section, where we describe the deep learning models used in this research along with the techniques for improving training time such as transfer learning. We also describe the data and the data and analysis procedures used in our study such as data augmentation to mitigate against imbalance. Next, we present the results of our analysis, including any findings. Finally, we discuss the implications of our results, conclude with a summary of our main findings, and suggest areas for future research.

## RELATED WORK

Research in the field of medical imaging, particularly in automating the screening and identification of TB from CXR images, has progressed significantly. Initial investigations explored traditional machine learning techniques, including support vector machines (SVM) [16, 17], decision trees [18, 19], random forests [20, 21], and XGBoost [22, 23], among others. However, recent advancements have shifted focus towards deep learning methods, such as Convolutional Neural Networks (CNNs), which have demonstrated promising results in image classification comparable to those of radiologists [13-15, 24]. Below, we review some of the recent studies that have utilised deep learning approaches for detecting (TB) in chest X-ray (CXR) images.

Hooda et al. [13] proposed a 19-layer convolution Neural Network (CNN) architecture for detecting TB, consisting of 7 convolutional layers, 7 Relu layers, 3 fully connected layers, and 2 dropouts layers. The model was trained on a dataset of 800 CXR images, each resized to 224×224 pixels. Utilizing the Adam optimizer, the study achieved notable results, with an overall accuracy of 94.73% and a validation accuracy of 82.09%. Although these results are impressive, the authors identified potential areas for further improvements. They suggested investigating the impacts of data augmentation and transfer learning on the model’s performance, highlighting avenues for future research enhancements and potential increases in accuracy.

Ojasvi et al. [25] developed a classification algorithm for CXR images of potential TB patients, aiming to improve upon existing models [26]. To mitigate against dataset imbalances and improve model reliability, they combined the NIH Chest X-ray Dataset, China-Shenzhen Chest X-ray Database, and Montgomery County Chest X-ray Database to train and fine-tune their model. By implementing coarse-to-fine transfer learning and extensive data augmentation techniques, they achieved a remarkable accuracy of 94.89% compared to the accuracy of 89.6% achieved by [26]. However, the study acknowledges the challenge of maintaining equivalent precision across CXR images obtained in varied settings as the model was specifically trained for the Chinese dataset.

Panicker et al. [27], introduced a novel two-stage detection method for TB bacilli, utilising image binarisation and CNN classification to analyze microscopic sputum smear images. The method was evaluated on a diverse dataset of 22 images, and the model demonstrated high effectiveness, achieving a recall rate of 97.13%, a precision of 78.4%, and an F-score of 86.76%. However, the study noted that the model’s ability to accurately detect overlapping bacilli was limited. In the same year, Stirenko et al. [28] explored the application of lung segmentation in CXR images and data augmentation to enhance TB detection from CXR images. Their study highlights the critical role of pre-processing, including lung segmentation and data augmentation, in addressing overfitting issues and improving the effectiveness of computer-aided diagnosis (CADx) systems in TB identification, particularly when working with limited datasets.

The study by Kazemzadeh et al. [15] developed a deep learning algorithm for detecting active pulmonary TB from CXR images. The algorithm was trained and validated on a dataset comprising 165,754 images from 22,284 subjects from 10 different countries. The algorithm’s performance was compared to that of 14 radiologists on datasets from four countries, including a cohort from a South African mining population. It achieved an AUC-ROC of 0.89, with superior sensitivity (88% vs. 75%, p¡0.05) and comparable specificity (79% vs. 84%) to radiologists, demonstrating its potential for TB screening in resource-limited settings. Another study by Nijiati et al. [29] utilized a 3D ResNet-50 CNN architecture to differentiate active from non-active pulmonary TB using CT images. This study, similar to that of Kazemzadeh et al. [15], reported high diagnostic accuracy and efficiency, outperforming conventional radiological methods in terms of speed and precision.

In their 2019 study, Meraj et al. [30] used CNN architectures such as VGG-16, VGG-19, ResNet50, and GoogLeNet to automate the detection of TB manifestations in CXRs, utilizing two public TB image datasets [31]. Their findings showed that the VGG-16 model outperformed other architectures in terms of accuracy and AUC-ROC. However, the study was limited by its reliance on small and unbalanced datasets, raising questions about the generalizability of the results. In contrast, our research builds upon and extends the work of Meraj et al. [30] by incorporating a larger and more diverse dataset. We also explore the diagnostic capabilities of more advanced CNN architectures, including ResNet101, ResNet152, and Inception-V2, to assess their effectiveness in TB detection. This approach aims to provide a more comprehensive understanding of how recent deep learning advancements can be leveraged for more accurate TB diagnosis in varied clinical settings.

## MATERIALS AND METHODS

In this section, we provide a comprehensive overview of the methodologies used in our study, including the dataset and preprocessing, data normalization, data augmentation, the application of transfer learning me hods, the architecture of CNNs utilized, and the evaluation metrics adopted to assess the performance of the models.

### Implementation Overview

The implementation framework illustrated in Figure 1 starts with the acquisition of a well-defined dataset, followed by comprehensive data pre-processing, which includes data augmentation, resizing, normalizatio, and partitioning into training, validation, and test sets. Subsequently, we embark on the development of various deep learning models. These models undergo extensive training and evaluation against different hyperparameters and evaluation metrics, to accurately predict and classify CXR images into positive or negative cases of TB.

**Figure 1.**
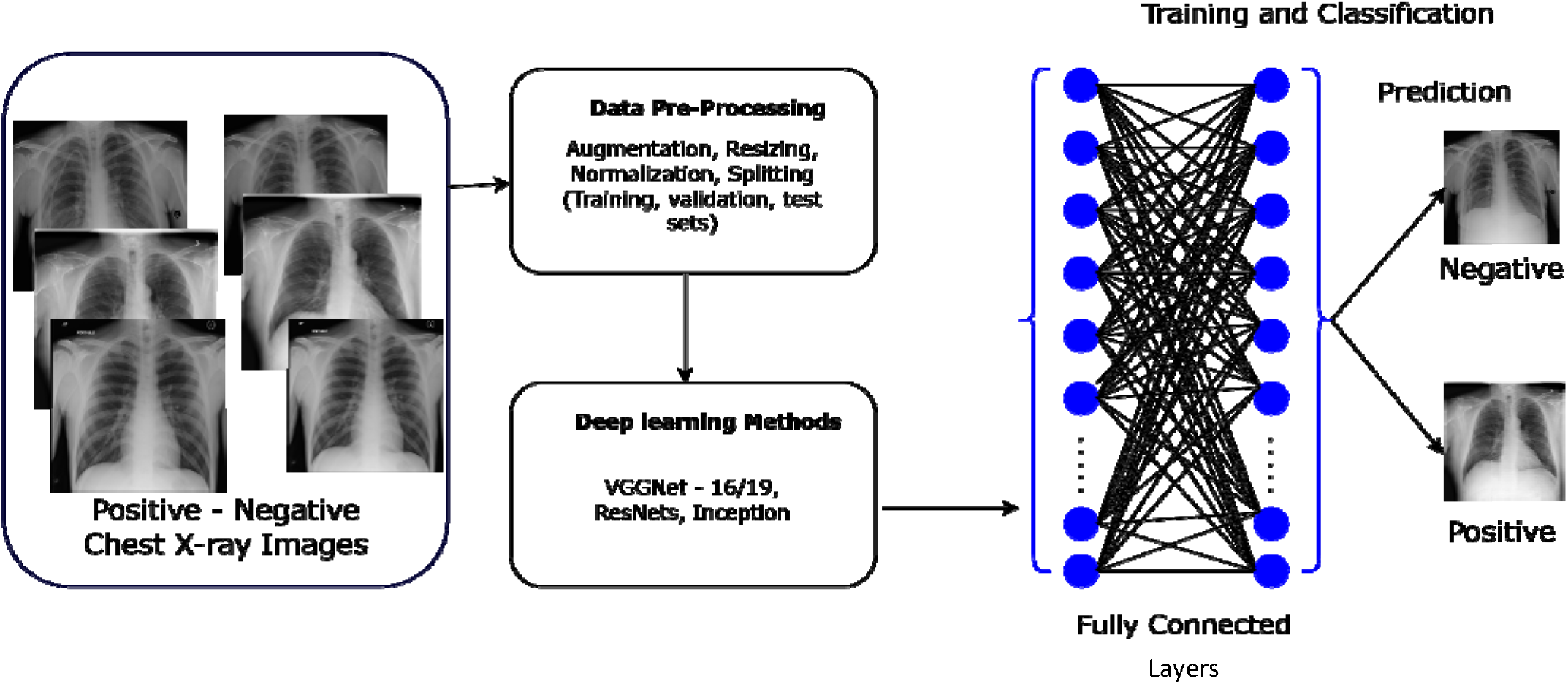
The implementation flow of the deep learning classification methodology

### Dataset

The dataset utilized in this research comprises 4,200 CXR images sourced from a public Kaggle data repository. The dataset was compiled through a collaborative effort between researchers from Qatar University (Doha, Qatar), the University of Dhaka (Bangladesh), and collaborators from Malaysia. They worked closely with medical professionals from the Hamad Medical Corporation (Doha, Qatar) and various healthcare institutions in Bangladesh. The dataset consists of 700 CXRs images indicative of TB and 3,500 CXRs images classified as normal, with all images having a resolution of 512×512 pixels [32]. This composition provides a substantial foundation for evaluating the effectiveness of CNN models in the detection of TB from CXR images. Figure 2 presents some of the images from the dataset.

**Figure 2.**
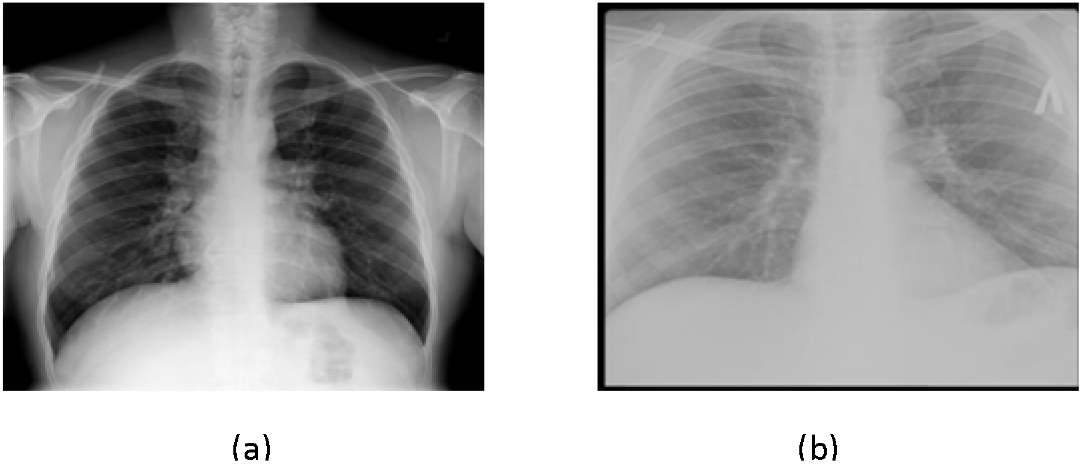
The CXR sample images (a) a TB-Negative (b) TB-positive

### Pre-Processing

To optimize the performance and efficiency of our models, we implemented key pre-processing techniques, specifically data normalization and augmentation, prior to training the models.

#### Data Normalization

In the pre-processing stage of image analysis, normalization is a critical step to standardize the input data, facilitating the model’s learning process. This study applies normalization to CXR images, which initially possess pixel intensity values in the range of 0 to 255, common for grayscale images [33]. The goal of normalization is to adjust these intensity values to a standardized scale that improves computational efficiency and model convergence during training. The normalization process is mathematically represented as follows:

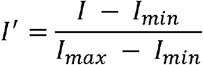

where:

- *I* represent the original pixel intensity of the image,
- *I*_*min*_ and *I*_*max*_ are the minimum and maximum possible intensity values in the original image, respectively,
- *I*^⍰^ is the normalized pixel intensity.

For grayscale images, *I*_*min*_ = 0 and *I*_*max*_ = 255. This equation effectively rescales the pixel intensity values to the range [0,1], making the input data more suitable for processing by the neural network layers. This normalization technique is advantageous because it ensures that each input parameter (pixel, in this case) contributes equally to the analysis, preventing features with initially larger ranges from dominating the learning process [34]. It also helps to stabilize the gradient descent optimization algorithm by maintaining a consistent scale for all gradients [35].

#### Data Augmentation

Data Augmentation represents a powerful regularization strategy designed to artificially increase the dataset through label-preserving transformations, thereby incorporating more invariant examples into the training set [36]. This approach, characterized by its computational efficiency, has been previously used to reduce overfitting when training CNNs, such as in the ImageNet Large-Scale Visual Recognition Challenge (ILSVRC), where it contributed to achieving state-of-the-art results [37]. This method enhances the robustness and generalizability of deep learning models by exposing them to a wider array of variations, simulating real-world variability. In our study, to address the imbalance between TB-positive and TB-negative images and to introduce different variations, we randomly augmented 30% of the TB-positive images and 5% of TB-negative images. The data augmentation techniques applied included random rotation within a range of 0 to 60 degrees, random width and height shifts of up to 0.2 times the image size, and random zooming of up to 0.2 times the original size, alongside horizontal and vertical flipping. To manage the newly created pixels from such transformations, a “fill mode” strategy was employed, ensuring integrity and consistency in the augmented images. These augmentations were performed using Keras’s ImageDataGenerator, a comprehensive data augmentation suite [38]. Figure 3 shows a sample of real images and their corresponding augmented outputs.

**Figure 3.**
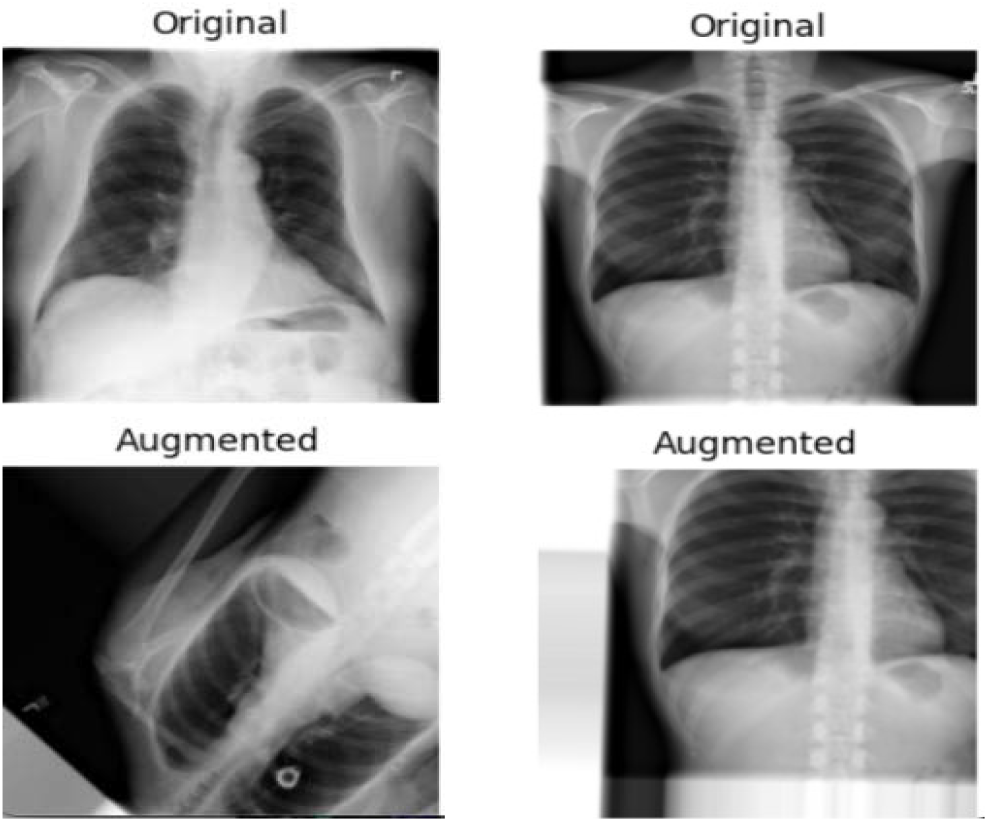
Sample of real and corresponding augmented images

### Transfer Learning

Transfer Learning is a machine learning technique where a model developed for a specific task is repurposed as the starting point for a model on a second, related task [39]. This technique leverages the knowledge gained during the initial training phase in one domain to enhance learning in another, potentially unrelated domain. It operates under the principle that information learned in one context can be exploited to accelerate or improve the optimization process in another, essentially allowing for the transfer of learned features and patterns across different but related problems [39]. This approach is particularly beneficial in situations where the dataset for the second task is too small to train a deep learning model from scratch or when computational resources are limited, offering a pathway to achieve high model performance with relatively less data and training time.

In this study, we propose an implementation that capitalizes on the transfer learning paradigm by utilizing pre-trained models such as Inception-v3, ResNet (50, 101, 152), and VGG (16, 19), which were initially trained on the ImageNet dataset [37]. This adaptation involves fine-tuning and customizing the models’ last layers to suit our classification task, effectively tailoring the robust, pre-learned representations of the ImageNet dataset to recognize and interpret the specific patterns and anomalies associated with TB in CXR images.

### CNN Architectures

In the next subsections, we provide a brief description of the VGG and ResNet families of CNN architectures as well as the Inception ResNet architecture that are considered in this study.

#### VGGNet

Introduced by Simonyan and Zisserman from the University of Oxford’s Visual Geometry Group in 2014, the VGGNet architecture marked a significant milestone in the field of deep learning [43]. Known for its outstanding performance in the ImageNet Large Scale Visual Recognition Challenge (ILSVRC) of that year, VGGNet is characterized by its use of 3×3 filters in all convolutional layers, simulating the effects of larger receptive fields. This architecture is available in two variants, VGG16 and VGG19, differing in depth and the number of layers, with VGG19 being the deeper model.

In our research, we utilized both the VGG16 and VGG19 architectures to train models on datasets consisting of solely real CXR images, and a combination of augmented and real images. This approach aimed to assess the impact of incorporating augmented images on the performance of these two architectures. Images were resized to 256×256 pixels before being input into the networks. We extended the architectures by adding a flattening layer, followed by a dense layer of 512 neurons with a *relu* activation function and a dropout layer with a dropout rate of 0.2 to mitigate overfitting. A softmax activation function was used in the output layer for binary classification. We employed the *Adam* optimizer with the *binary crossentropy* loss function for optimisation. The training was conducted over 15 epochs with a batch size of 32 for both models. This rigorous approach ensured both architectures could classify between TB-positive and TB-negative CXR images accurately.

#### ResNet

[41] introduced the deep residual network (ResNet) architecture in their 2016 seminal paper. This architecture greatly improved the performance of deep neural networks and went on to win the COCO object detection challenge and the 2015 ImageNet Large Scale Visual Recognition Challenge (ILSVRC). To date, several variants of the ResNet architecture exist, including ResNet50, ResNet101, and ResNet152, which vary in depth and number of layers. ResNet architectures are very deep models [41, 44]. The core idea behind ResNet is the use of residual connections, also known as shortcuts, which bypass one or more layers. By resolving the vanishing gradient issue, these shortcuts maintain the gradient flow across the network and facilitate the training of much deeper networks [41].

The CXR images in this study were classified using the ResNet50, ResNet101, and ResNet152 architectures. We added three more layers to the ResNet50 model, two, each with 256 units and one with 512 units, using batch normalization and ReLU activation in each layer. To reduce overfitting, dropout layers were added with dropout rates of 0.3, 0.25, and 0.2 respectively. The binary cross-entropy loss function was used to compile the model, while the Adam optimizer was used to optimise the model at a learning rate of 0.001. Two units with a softmax activation function made up the output layer, which classified the images as either TB-positive or TB-negative. Training for this model involved 16 batch sizes and 100 epochs.

ResNet101 was trained using the same settings as ResNet50, as preliminary training showed that the same parameter values used for ResNet50, also yielded optimal results for the ResNet101 architecture. For ResNet152, a selective fine-tuning approach was adopted where, only the last 10 layers of the network were trainable, enhancing the model’s focus on more feature-specific adjustments in the later stages of the network. This model shared the augmentation layers of ResNet50 but was trained for only 50 epochs, incorporating a learning rate scheduler, *ReduceLROnPlateau*, which adjusted the rate based on the validation loss with a factor of 0.1, patience of 5, and a minimum learning rate of 1×10^−6^, thereby optimizing the training dynamics. The details of the models’ configuration are shown in Table 1.

**Table 1:** Training Hyperparameters Of ResNet Models

| Hyperparameter | ResNet50 | ResNet101 | ResNet152 |
| --- | --- | --- | --- |
| Number of Layers | 53 (50 base + 3 extra) | 104 (101 base + 3 extra) | 155 (152 base + 3 extra) |
| Units per Layer | 256, 256, 512 | 256, 256, 512 | 256, 256, 512 |
| Activation | ReLU | ReLU | ReLU |
| Batch Normalization | Yes | Yes | Yes |
| Dropout Rate | 0.3, 0.25, 0.2 | 0.3, 0.25, 0.2 | 0.3, 0.25, 0.2 |
| Optimizer | Adam | Adam | Adam |
| Learning Rate | 0.001 | 0.001 | Variable (ReduceLROnPlateau) |
| Loss Function | Binary Crossentropy | Binary Crossentropy | Binary Crossentropy |
| Training Epochs | 100 | 100 | 50 |
| Batch Size | 16 | 16 | 16 |

#### Inception-ResNet

The Inception networks, introduced by Szegedy et al. [45], have greatly advanced the field of CNN as they have achieved state-of-the-art performance in a number of computer vision problems [45-47]. The original Inception V1, also known as GoogleNet, was first introduced in 2014 and won the ILSVRC of that year. The architecture introduced a novel approach, of using multiple convolutional filter sizes in parallel, allowing the network to capture various spatial features of different scales with improved utilization of computing resources [45].

In this study, we used Inception-ResNet V2 architecture, a hybrid model that combines the benefits of both the Inception and residual networks. This hybrid approach enables the architecture to learn more complex features with improved training stability and faster convergence [45]. The Inception-ResNet V2 also leverages residual connections to skip certain layers during training, which helps it improve gradient flow, accelerate training times, and reduce the likelihood of vanishing gradient problems in deep networks [48]. We selected Inception-ResNet V2 due to its demonstrated state-of-the-art results in several medical imaging tasks [47].

For our implementation, the Inception-ResNet V2 architecture was initialized with weights pre-trained on the ImageNet dataset. Similar to our approach with the ResNet152 model, all layers except the last ten were frozen to retain the pre-trained features from ImageNet. The last ten layers were set to be trainable, enabling the model to learn specific features from the CXR images. We added three new layers: two with 256 units each and one with 512 units, all using ReLU activations and Batch Normalization. Each of these layers was followed by Dropout layers with rates of 0.4, 0.35, and 0.3, respectively, to introduce non-linearity and reduce overfitting. The final output layer consisted of two units with a softmax activation function for binary classification. The model was then compiled using binary cross-entropy as the loss function and the Adam optimizer with a learning rate of 0.0001. Training was conducted for 50 epochs with a batch size of 16.

### Evaluation Metrics

The performance of the CNN architectures in classifying CXR images into TB-positive and TB-negative categories was assessed using several standard performance metrics including: accuracy, precision, recall, F1-score, and the Area Under the Receiver Operating Characteristic curve (AUC-ROC). Each metric provides unique insights into the model’s classification abilities, considering both the true and false predictions.

#### Accuracy

This metric measures the proportion of true positive and true negative results among the total number of cases examined:

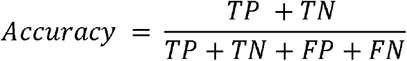

where:

- TP (True Positives): The number of TB-positive images that are correctly identified as TB-positive by the model
- TN (True Negatives): The number of TB-negative images that are correctly identified as TB-negative by the model
- FP (False Positives): The number of TB-negative images that are incorrectly identified as TB-positive by the model
- FN (False Negatives): The number of TB-positive images that are incorrectly identified as TB-negative by the model.

#### Precision

Also known as *positive predictive value*, precision is the ratio of correctly identified TB cases to all cases that were diagnosed as TB by the model. It measures the model’s accuracy in diagnosing a patient with TB when the model predicts the disease. High precision indicates a low rate of false TB diagnoses. Mathematically, it is defined as:

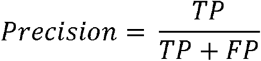

#### Recall

Recall, or sensitivity, is especially critical in medical diagnostics as it quantifies the model’s ability to correctly identify all actual TB cases. It represents the proportion of actual TB cases that were correctly identified by the model and aims to minimize the risk of missing a true TB case. It is computed as:

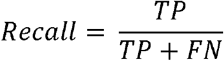

#### F1-Score

The F1-score is the harmonic mean of precision and recall, providing a single measure that balances both the false positives and false negatives. In TB diagnosis, it is particularly useful because it creates a balance between precision (minimizing false TB diagnoses) and recall (minimizing missed TB diagnoses), which is crucial for medical screening tests. It is defined as:

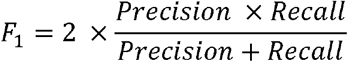

#### AUC-ROC

The Area Under the Receiver Operating Characteristic Curve (AUC-ROC) measures a model’s ability to discern between positive and negative classes. In the context of our problem, that specifically refers to distinguishing between TB-positive and TB-negative CXR images. The AUC-ROC curve is a plot of the true positive rate (TPR) against the false positive rate (FPR) at various threshold settings. The AUC-ROC provides an aggregated measure of the model’s performance across all classification thresholds, with a value of 1 representing a perfect model and a value of 0.5 representing a model with no discriminatory power. The approximate area under the receiver operating characteristic curve (AUC-ROC) is calculated by using the following formula:

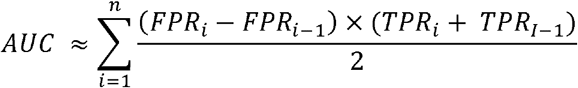

where *i* is the current data point or threshold, *FPR*_*i*_ and *TPR*_*i*_ are the false positive and true positive rates at the *i*-th threshold, respectively, and *n* is the number of data points or thresholds used to calculate the AUC-ROC. Each term in the sum represents the area of a trapezoid where (*FPR*_*i*_ *− FPR*_*i−1*_) is the base of the trapezoid and (*TPR*_*i*_ *+ TPR*_*i−*1_)/2 is the average height of the trapezoid. The formula calculates the AUC-ROC by summing the areas of trapezoids formed by connecting consecutive points on the AUC-ROC curve.

### Computational Environment

The implementation and findings of this study were based on utilizing the Keras 3.3.3 and TensorFlow 2.16.1 frameworks. The experiments were conducted on a single GPU MSI GL75 Leopard 10SFR laptop with 32GB of RAM and an 8GB NVIDIA GEFORCE RTX 2070 GDDR6 card. The system operated using the CUDA 12.1 and cuDNN SDK 8.7.0 platforms to ensure efficient GPU acceleration and deep learning model training.

## RESULTS

The study aimed to analyze and compare the performance of various CNN architectures, including VGG16, VGG19, ResNet50, ResNet101, ResNet152, and Inception-ResNet-V2, in classifying CXR images as either TB-positive or TB-negative. Additionally, we also investigated whether data augmentation could further improve the classification performance of these models by comparing the performance of models trained on only real images versus those trained on a combination of real and augmented data. We went further to examine the training time and the number of parameters for each architecture to understand the computational efficiency and resource demands for each model. This analysis is important for practical implementation, particularly in resource-constrained settings where training time and computational costs are significant considerations. By evaluating these parameters, we aimed to identify models that not only perform well but also offer a balanced trade-off between accuracy and efficiency, making them suitable for real-world applications in diverse healthcare environments.

Table 2 presents the models’ performance evaluated across accuracy, precision, recall, and F-score for models trained on real images and those trained on a combination of real and augmented data. Figure 4 shows the performance of these models when evaluated using the AUC-ROC score metric. It is observed that the VGG16 outperformed all other architectures across all metrics, with an accuracy of 99.4%, precision of 97.9%, recall of 98.6%, F1-Score of 98.3%, and AUC of 98.25%. Its performance was superior consistently irrespective of whether the models were trained with or without data augmentation.

**Table 2:** Evaluation of CNN architectures across key evaluation metrics: This table summarizes the performance of various CNN architectures according to precision, recall, and F1-Score. Models marked with an asterisk (*) were trained using a combination of real and augmented data, showcasing the impact of data augmentation on model performance.

| Architecture | Accuracy | Precision | Recall | F1-Score |
| --- | --- | --- | --- | --- |
| VGG16 | 99.4 | 97.9 | 98.6 | 98.3 |
| VGG16* | 99.3 | 96.6 | 99.3 | 97.9 |
| VGG19 | 99.2 | 96.6 | 98.6 | 97.6 |
| VGG19* | 99.2 | 96.6 | 98.6 | 97.6 |
| ResNet50 | 96.1 | 81.3 | 96.9 | 88.4 |
| ResNet50* | 89.0 | 97.5 | 30.0 | 45.9 |
| ResNet101 | 96.9 | 94.8 | 84.6 | 89.3 |
| ResNet101* | 97.3 | 92.1 | 90.0 | 91.1 |
| ResNet152 | 97.9 | 93.6 | 93.6 | 93.6 |
| ResNet152* | 97.5 | 87.6 | 96.6 | 92.1 |
| Inception ResNet-v2 | 99.0 | 95.9 | 98.6 | 97.2 |
| Inception ResNet-v2* | 99.2 | 97.2 | 97.9 | 97.5 |

**Figure 4.**
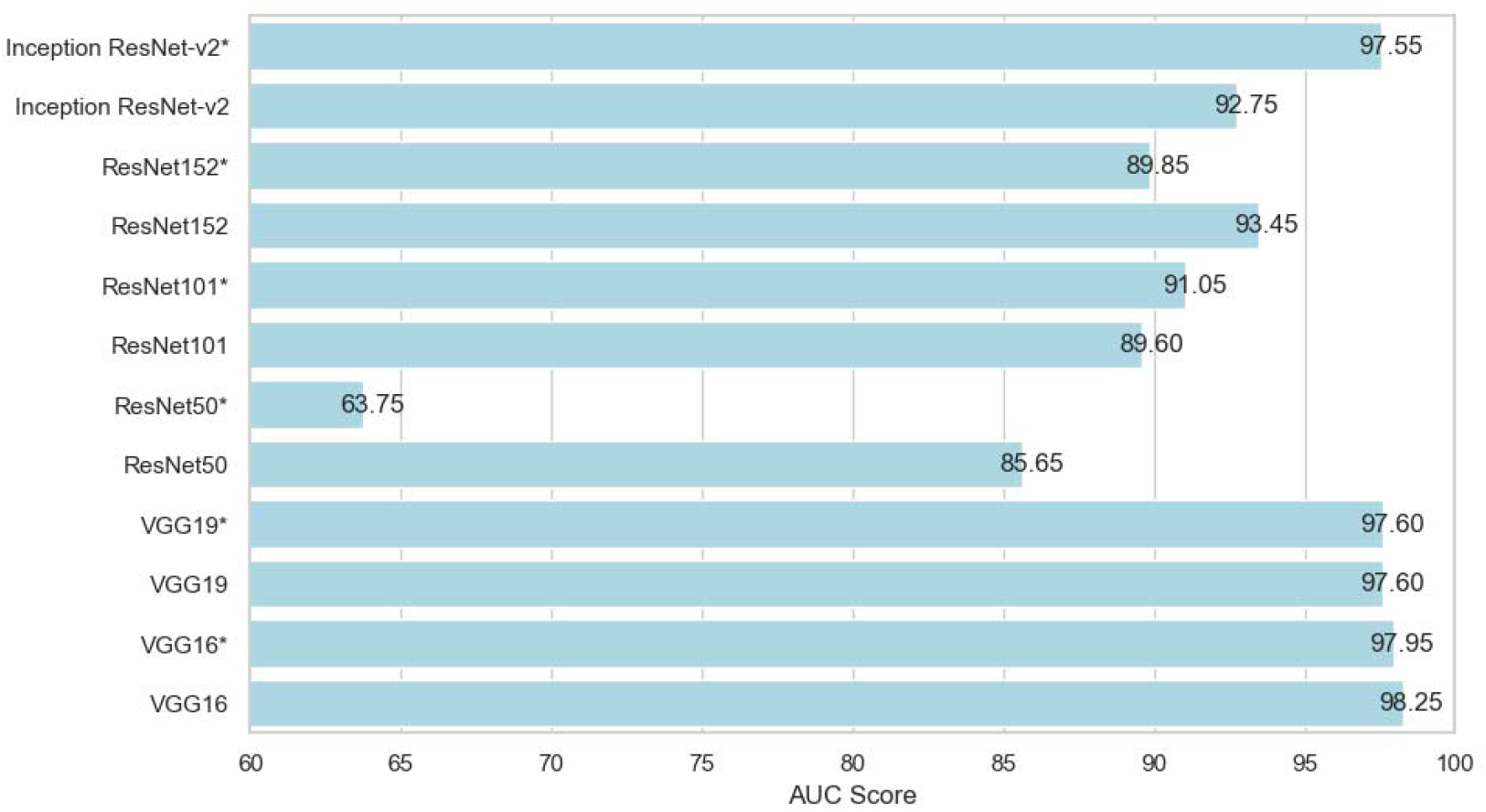
The models’ area under a curve scores

Surprisingly, increasing the dataset size through data augmentation did not correspond with an increase in the performance of the models across all architectures, as seen in Table 2. This was also observed in other models such as Resnet50, where when augmented data was included, the AUC-ROC score dropped significantly from 85.65% to 63.75% as shown in Figure 4.

### Training Time

We also tracked each model’s training time with a combination of data augmentation and real images versus training with only real images, as shown in Figure 5. As expected, training with data augmentation requires more time due to the increased size of the dataset. For example, training the ResNet152 with data augmentation took 356.6 minutes whereas training without augmentation took 345.7 minutes. This observation highlights the trade-off between longer training times and the potential benefits of data augmentation. However, data augmentation did not improve performance in our case, indicating that the additional training time did not translate into better model generalization.

**Figure 5.**
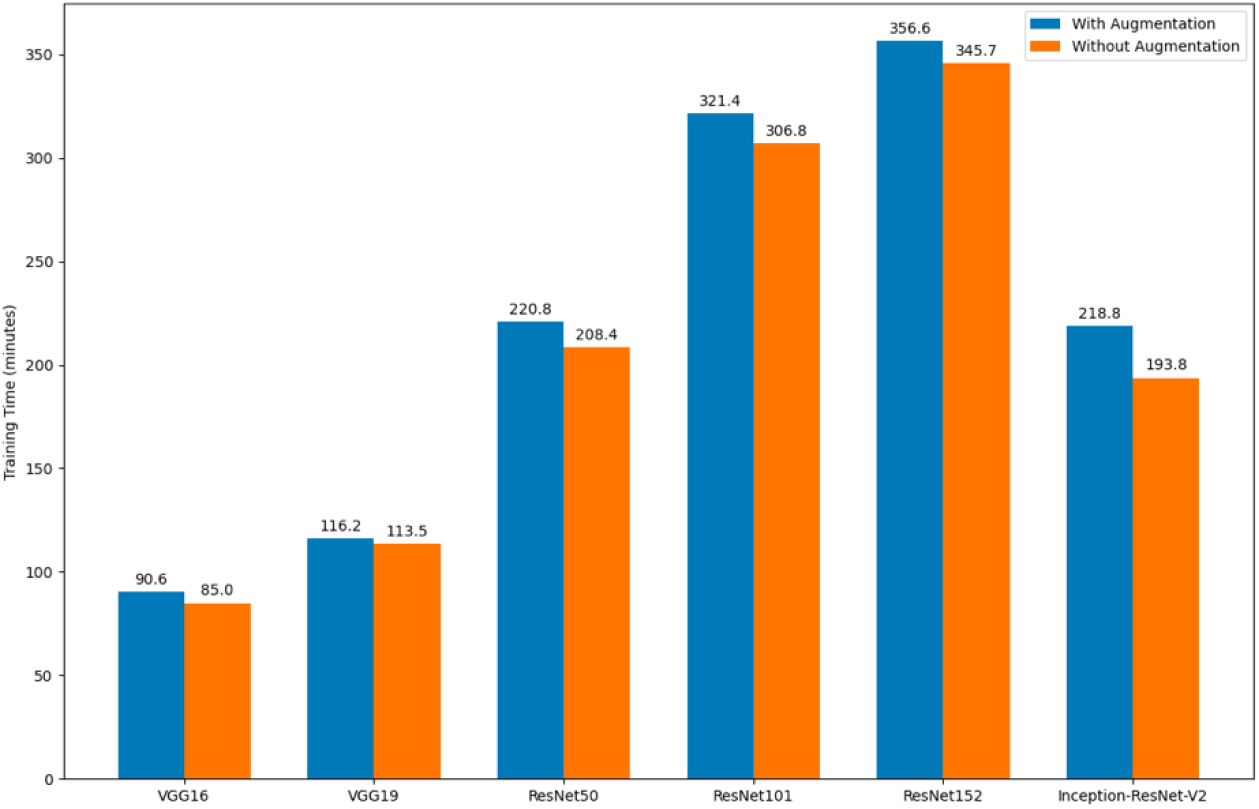
Training time for the models

### Model Parameters

In addition to our analysis, We provide a detailed breakdown of the parameter count for each model used in our study, as shown in Figure 6. The number of parameters in a model reflects its complexity and capacity to learn from data. Consequently, it has a direct impact on both training time and the computational resources required, influencing the model’s overall efficiency and scalability.

**Figure 6.**
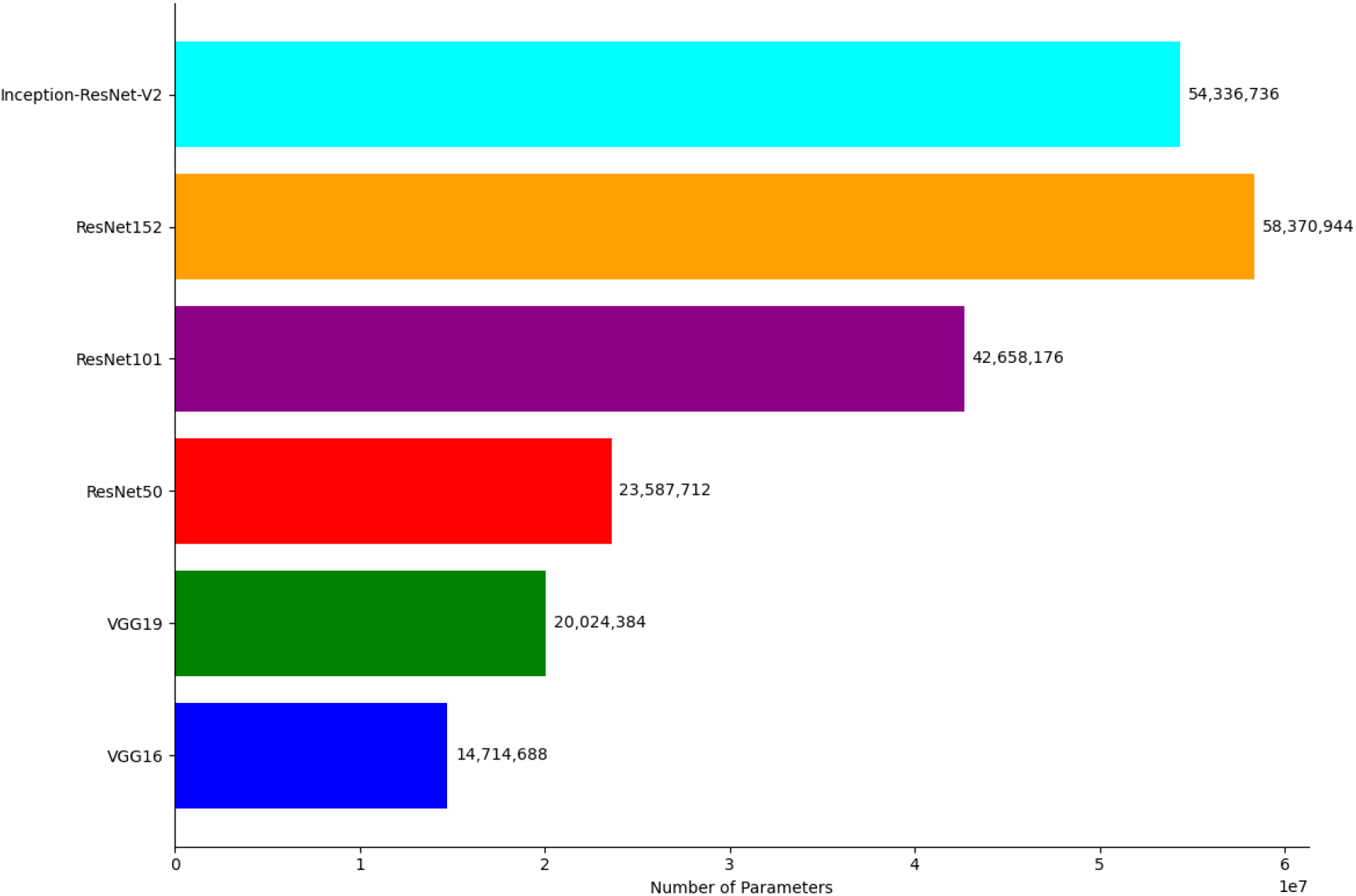
Parameters of each model

## DISCUSSION

The findings from this study provide significant insights into the performance and efficiency of several CNN architectures in the classification of CXR images for TB detection. The architectures evaluated included VGG16, VGG19, ResNet50, ResNet101, ResNet152, and Inception-ResNet-V2. Of these, the VGG16 consistently achieved the highest performance across all metrics such as: accuracy, precision, recall, and F1-score. This consistent performance suggests that VGG16 effectively captures the necessary features for distinguishing between TB-positive and TB-negative CXR images, even with fewer parameters compared to the deeper models.

The findings also highlight the fact that, while data augmentation is often used to improve the performance of CNN models by expanding the dataset and introducing variability, it does not necessarily lead to performance improvements if the base dataset already provides sufficient diversity for training. In our study, the original dataset appeared robust enough, and the addition of augmented data did not enhance model performance. This suggests that the benefits of data augmentation are context-dependent and may not always be necessary or effective when the existing dataset is already well-suited for the task.

It is commonly observed in several studies that models with a higher number of parameters, such as ResNet152 and InceptionResNet-V2, are capable of capturing more deep patterns in the data [41, 45]. However, this comes at the cost of requiring more computational resources and longer training times. Interestingly, in our study, despite having fewer parameters, VGG16 outperformed the more complex models. This suggests that for our specific task of classifying CXR images into TB-positive and Tb-negative categories, VGG16 efficiently captured the relevant features without necessitating excessive complexity. This finding highlights the importance of selecting the appropriate model architecture based on the specific characteristics and requirements of the task at hand, rather than simply opting for the model with the most parameters. This result also aligns with the principle that simpler models can often perform competitively when they are well-matched to the data and the problem domain [43].

## CONCLUSION

This study presents a comprehensive evaluation of several CNN architectures - VGG16, VGG19, ResNet50, ResNet101, ResNet152, and Inception-ResNet-V2 — in classifying CXR images as either TB-positive or TB-negative. The findings showed that the VGG16 architecture consistently outperformed the other models across all the evaluation metrics, achieving superior performance despite having fewer parameters compared to the more complex architectures such as ResNet152 and InceptionResNet-V2.

Our results also showed limited benefits of data augmentation in this context, suggesting that the original dataset provided sufficient diversity for effective training. This highlights the importance of context in the application of data augmentation techniques. Furthermore, the study demonstrated the significant trade-offs between model complexity, training time, and performance. Models with more parameters, such as ResNet152 and Inception-ResNet-V2, required longer training times and more computational resources without corresponding improvements in classification performance across all the evaluation metrics. This result continues to emphasize the need for careful selection of model architecture based on the specific characteristics and requirements of the task at hand. Overall, our research contributes to the growing body of evidence supporting the effectiveness of deep learning models in medical image classification and provides insights into optimizing these models for TB detection in CXR images.

## Data Availability

The data used in this study can be accessed from Kaggle via the following link: https://www.kaggle.com/datasets/tawsifurrahman/tuberculosis-tb-chest-xray-dataset

## ACKNOWLEDGMENT

We would like to extend our sincere gratitude to the team of researchers from Qatar University, Doha, Qatar, and the University of Dhaka, Bangladesh, along with their collaborators from Malaysia and medical doctors from Hamad Medical Corporation and various healthcare institutions in Bangladesh, for creating and sharing the CXR image database for TB. Their effort in compiling this comprehensive dataset has significantly contributed to our research. We are grateful for their contributions and their dedication to advancing TB diagnosis and treatment through the provision of this valuable public dataset.

## ETHICS APPROVAL

Since this study utilized a publicly available dataset from Kaggle, it did not require any ethical approval.

